# 15 Years of Longitudinal Genetic, Clinical, Cognitive, Imaging, and Biochemical Measures in DIAN

**DOI:** 10.1101/2024.08.08.24311689

**Authors:** Alisha J. Daniels, Eric McDade, Jorge J. Llibre-Guerra, Chengjie Xiong, Richard J. Perrin, Laura Ibanez, Charlene Supnet-Bell, Carlos Cruchaga, Alison Goate, Alan E. Renton, Tammie L.S. Benzinger, Brian A. Gordon, Jason Hassenstab, Celeste Karch, Brent Popp, Allan Levey, John Morris, Virginia Buckles, Ricardo F. Allegri, Patricio Chrem, Sarah B. Berman, Jasmeer P. Chhatwal, Martin R. Farlow, Nick C. Fox, Gregory S. Day, Takeshi Ikeuchi, Mathias Jucker, Jae-Hong Lee, Johannes Levin, Francisco Lopera, Leonel Takada, Ana Luisa Sosa, Ralph Martins, Hiroshi Mori, James M. Noble, Stephen Salloway, Edward Huey, Pedro Rosa-Neto, Raquel Sánchez-Valle, Peter R. Schofield, Jee Hoon Roh, Randall J. Bateman, the Dominantly Inherited Alzheimer Network

## Abstract

This manuscript describes and summarizes the Dominantly Inherited Alzheimer Network Observational Study (DIAN Obs), highlighting the wealth of longitudinal data, samples, and results from this human cohort study of brain aging and a rare monogenic form of Alzheimer’s disease (AD). DIAN Obs is an international collaborative longitudinal study initiated in 2008 with support from the National Institute on Aging (NIA), designed to obtain comprehensive and uniform data on brain biology and function in individuals at risk for autosomal dominant AD (ADAD). ADAD gene mutations in the amyloid protein precursor (*APP*), presenilin 1 (*PSEN1*), or presenilin 2 (*PSEN2*) genes are deterministic causes of ADAD, with virtually full penetrance, and a predictable age at symptomatic onset. Data and specimens collected are derived from full clinical assessments, including neurologic and physical examinations, extensive cognitive batteries, structural and functional neuro-imaging, amyloid and tau pathological measures using positron emission tomography (PET), flurordeoxyglucose (FDG) PET, cerebrospinal fluid and blood collection (plasma, serum, and whole blood), extensive genetic and multi-omic analyses, and brain donation upon death. This comprehensive evaluation of the human nervous system is performed longitudinally in both mutation carriers and family non-carriers, providing one of the deepest and broadest evaluations of the human brain across decades and through AD progression. These extensive data sets and samples are available for researchers to address scientific questions on the human brain, aging, and AD.

## Background and Summary

The vast majority of AD dementia is sporadic and generally occurs in older ages, but a small proportion (less than 1%) of AD dementia is caused by mutations in the Aβ precursor protein (*APP*), presenilin 1 (*PSEN1*), or presenilin 2 (*PSEN2*) genes with almost 100% penetrance, generally at predictable younger ages. This form of AD is known as autosomal dominant AD (ADAD). ADAD is thought to have an underlying pathogenic process similar to that of the more common sporadic AD and thus may hold the key to understanding the pathogenesis of AD and identification of effective treatments for both ADAD and sporadic AD. To leverage this population’s potential for AD research, the DIAN Obs was established in 2008. It aims to track individuals from families with known ADAD mutations, employing a wide array of cognitive assessments and biomarker tests. Since inception time, the DIAN Obs has amassed a substantial repository of clinical and biomarker data and samples, facilitating a deeper understanding of ADAD’s natural history.

The DIAN Obs cohort has been longitudinally followed for over 15 years and is amongst the most deeply phenotyped cohorts of brain aging, function, and AD measures in predominantly 18 to 55-year-old people. The inaugural year of DIAN Obs included ten performance sites in three countries (US, UK, and Australia) with English as the language for all initial sites. The continued success of DIAN Obs has since grown to twenty-three performance sites in eleven countries and supporting seven languages: English, Spanish, German, Japanese, Korean, French, and Portuguese (Figure 1 and Supplement Table 1). A total of 664 participants have enrolled in DIAN Obs; currently the study has 314 currently active participants.

**Figure 1:**
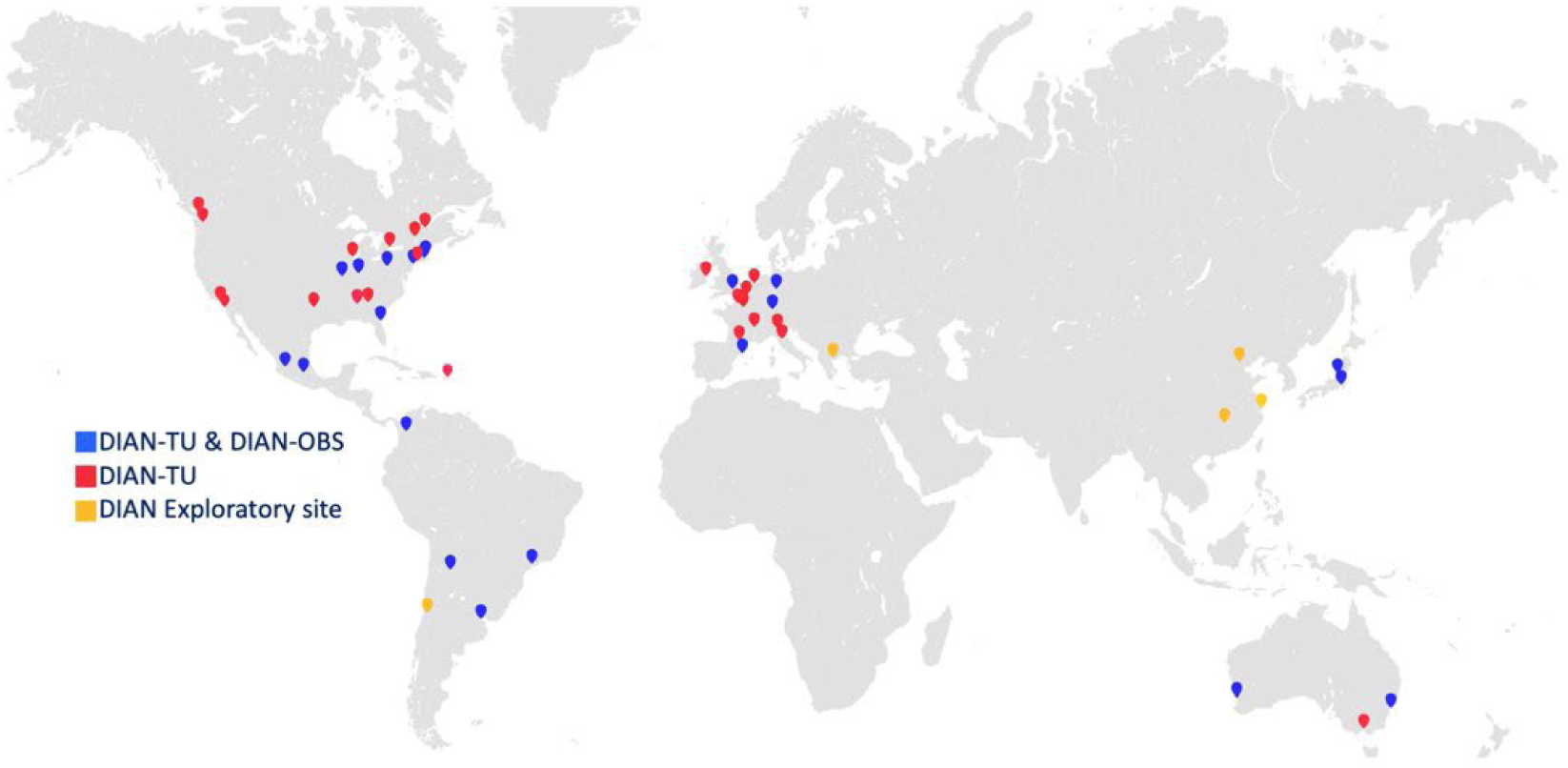
DIAN Obs and DIAN-TU Map

DIAN Obs is the key scientific and discovery study for ADAD and also provides natural history information for the DIAN-Trials Unit (DIAN-TU), which is the therapeutic and target validation platform for treatment and prevention trials. The DIAN-TU [<u>dian.wustl.edu/our-research/clinical-trial/</u>]^6–7^ is a global research effort established in 2012 to design and conduct clinical trials for the prevention or treatment of ADAD. Data from DIAN Obs and DIAN-TU studies were designed to be conducted together, with nearly identical protocols, including cognitive and clinical assessments, biomarker measures, and quality control.

Participant outreach, recruitment, and retention for both DIAN Obs and DIAN-TU are facilitated via the international DIAN Expanded Registry (DIAN EXR) [<u>dian.wustl.edu/our-research/registry</u>], established in 2012 for individuals who are or may be affected by ADAD. DIAN EXR serves as a collaborative research effort not only to facilitate study referral to DIAN Obs and DIAN-TU, but also to support educational and outreach activities with ADAD family members.

DIAN Obs and DIAN-TU have distinct purposes with differing eligibility requirements and site locations allowing the DIAN EXR to act as a central mechanism for navigation and referral of interested registrants to potential research opportunities (Figure 2).

**Figure 2:**
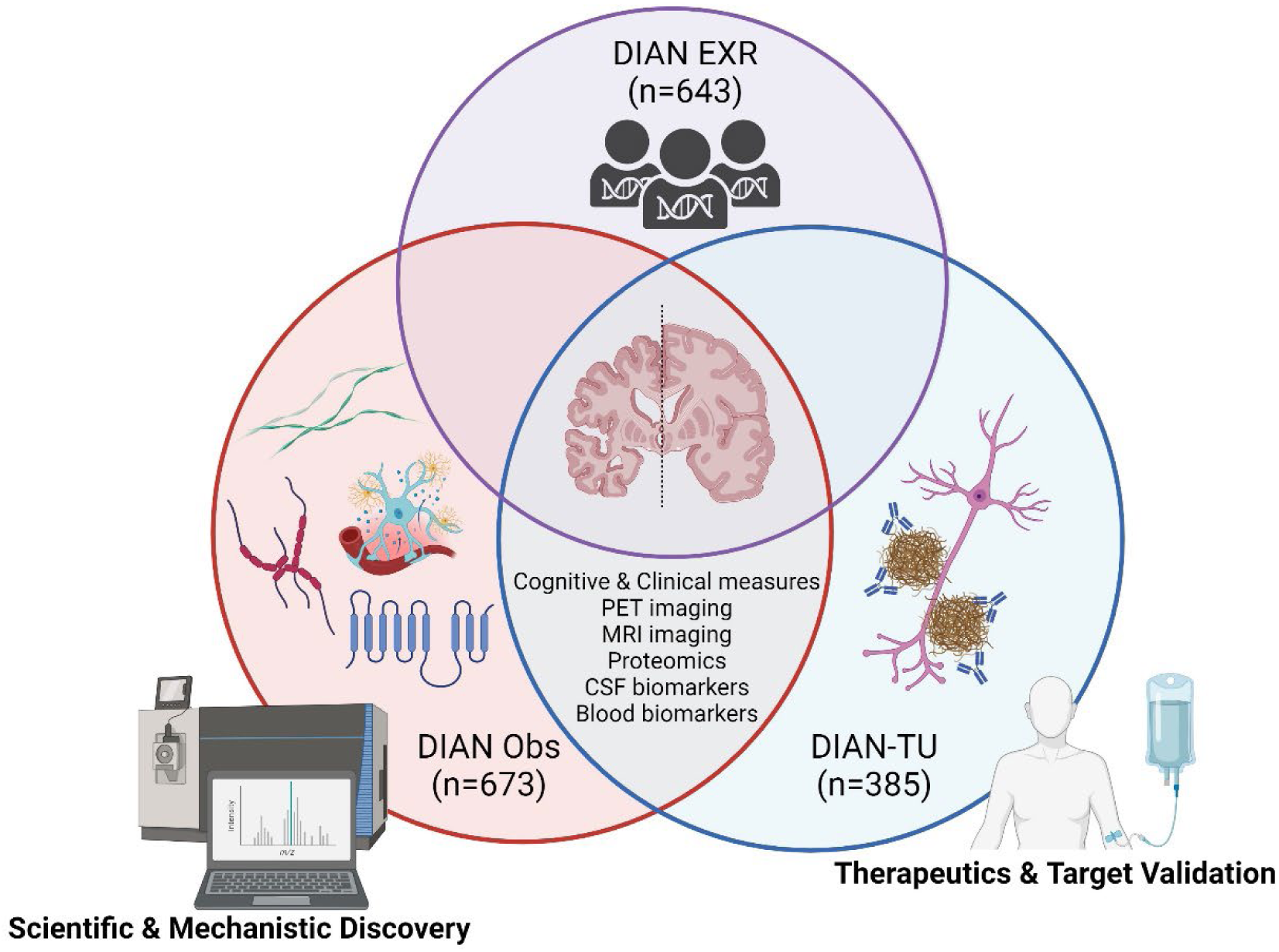
Coordination of DIAN Obs, DIAN-TU, and DIAN EXR

The main scientific hypotheses of DIAN Obs aim to address fundamental questions regarding the pathophysiology of AD. First, AD biomarker changes identify mutation carriers (MCs) many years before symptomatic AD develops, thus supporting the concept of preclinical AD. Second, the initial biomarker changes in the preclinical stage of ADAD involve Aβ42, followed by changes related to neurodegeneration, followed by cognitive decline. Third, the clinical and neuropathological phenotypes of ADAD will reflect those of “sporadic” late onset AD (LOAD). DIAN Obs emphasizes longitudinal data as it provides more accurate and precise information on the magnitude and rate of change for biofluid and imaging biomarkers throughout the preclinical stage of ADAD.

DIAN Obs has provided seminal advances in the understanding of brain health, the onset and progression of ADAD, and how this compares to the more common sporadic AD (Figure 3). In 2012, DIAN Obs described a comprehensive order of clinical, cognitive, imaging, and biomarker changes that occur across time span two decades before and a decade after symptom onset. Amyloid plaques continuously accumulate for 15-20 years before symptom onset, defining an amyloid growth phase,^1–4^ while tau tangles appear and accumulate in the transition to and during the symptomatic phase.^5^ A series of biochemical changes in CSF and blood begin with amyloid-beta 42/40 decreasing, followed closely by p-tau217/181/231 associated with amyloid plaques, then p-tau205, neurofilament light chain (NfL), and total tau increasing before the appearance of tangles. Cortical hypometabolism begins five to ten years before symptom onset, and cortical atrophy five years before symptom onset. Finally, changes in cognitive performance are detected several years before onset.

**Figure 3:**
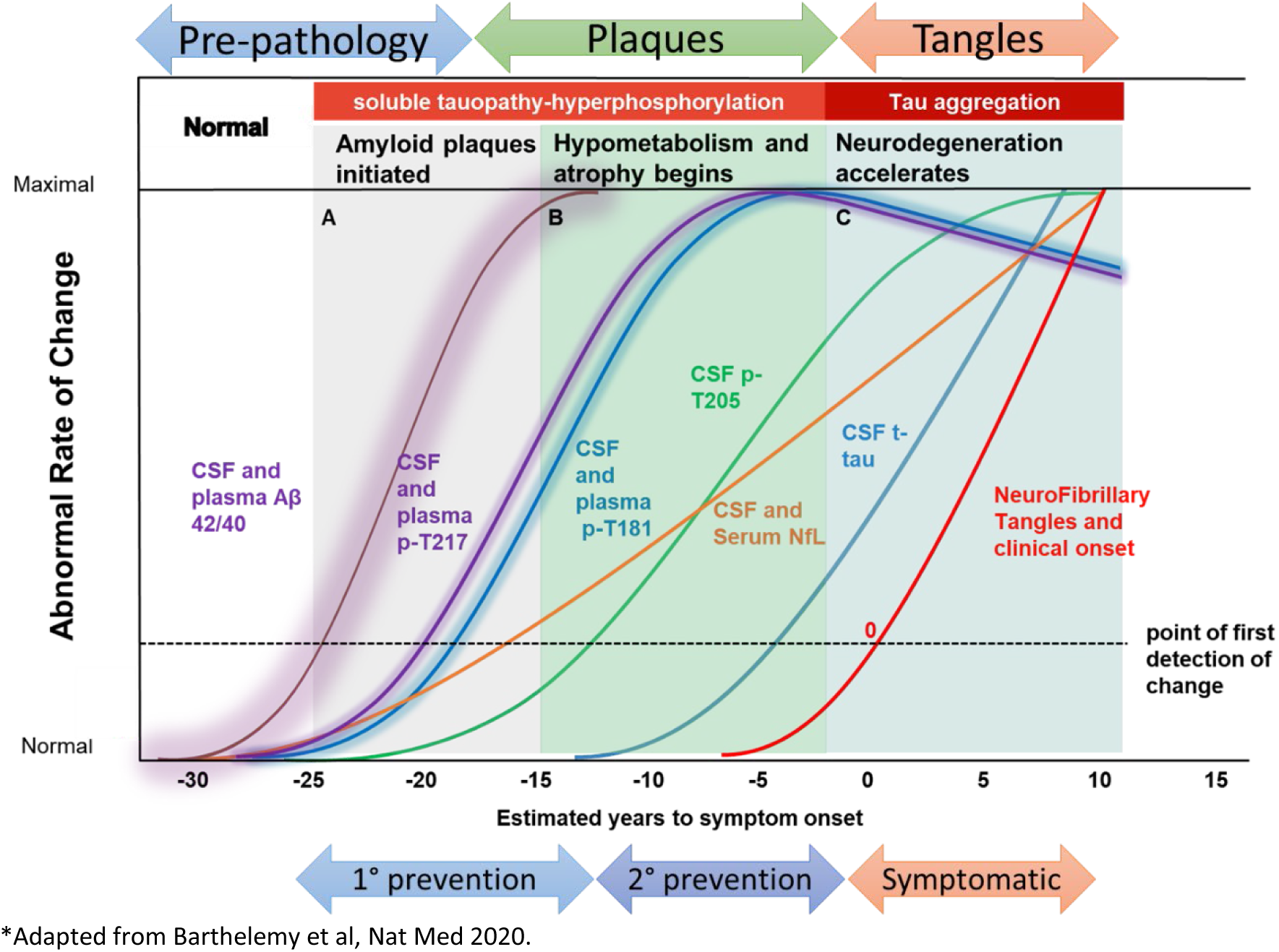
ADAD Onset & Progression compares to Sporadic AD

DIAN-TU implements therapeutic trials in a trial platform with the goal to slow, delay, or prevent dementia in the ADAD population.^6,7^ DIAN Obs and DIAN-TU implement harmonization efforts to ensure the combination and comparability of DIAN-TU and DIAN Obs protocols including International Council for Harmonisation Good Clinical Practice (ICH GCP) guideline compliance. These harmonization efforts increase longitudinal data and sample resources that can be combined across DIAN Obs and DIAN-TU. The Biostatistics Core enables the ability to link coded participant data across the DIAN Obs and DIAN-TU, enabling combining and comparing across the studies. Key measures harmonized include electronic capture of clinical and cognitive assessments, sample collection protocols, and imaging (MRI and PET).

## Methods

### A. Overview

Washington University in St. Louis is the recipient of a U19 grant from the NIA and serves as the DIAN Obs Coordinating Center which oversees both scientific and administrative center and as a performance site. The DIAN Obs Coordinating Center consists of eight Cores: Administration, Clinical, Genetics, Cognition, Imaging, Biomarker, Neuropathology, and Biostatistics and three scientific Projects: Amyloid-β, Tau, and Novel Mechanisms. All performance sites have access to adequate numbers of potential DIAN Obs participants and the resources and capabilities to conduct all elements of the DIAN Obs protocol.

### B. Institutional Review Board & Consenting Statement

The Institutional Review Board (IRB) of Washington University School of Medicine in St. Louis approved the study with the IRB ID# 201106339, and research was performed in accordance with the approved protocols. Participants review and discuss the consent form with performance site study team prior to being asked to sign the informed consent for study participation. A copy of the consent is provided to participants and the original is maintained in the participant’s research record.

### C. Administration Core

The DIAN Obs Administration Core provides oversight and management of the DIAN Obs project including: coordinating activities of the other Cores, scientific research Projects, and subcontractors; managing and supporting the DIAN Obs Steering Committee; seeking and facilitating feedback from the External Advisory Committee; interacting with the NIA liaisons; and managing activation, maintenance, and data collection of all performance sites.

The Administration Core has established a web-based system to support data and tissue resource dissemination to investigators. The DIAN Obs data and biospecimen application form is available on the DIAN website (<u>dian.wustl.edu</u>). All requests are reviewed by relevant Core Leaders, the Study Director, and DIAN Obs Steering Committee. Upon receiving request approval, with appropriate institutional review board (IRB)/institutional ethics committee (IEC) approvals and data and tissue sharing agreements, data and/or biospecimens are shared with support from the related Cores and Biostatistics Core. The DIAN Obs Data and Tissue Sharing, Notifications, Publications, and Authorship Policies [<u>DIAN</u> <u>Publication Policy</u>] govern the sharing of DIAN Obs resources and guidelines for publications. As of December 2022, DIAN Obs has received 288 data requests and 114 tissue requests, with 231 and 85 requests fulfilled respectively. An overview of resources that may be requested is provided in Figure 4.

**Figure 4:**
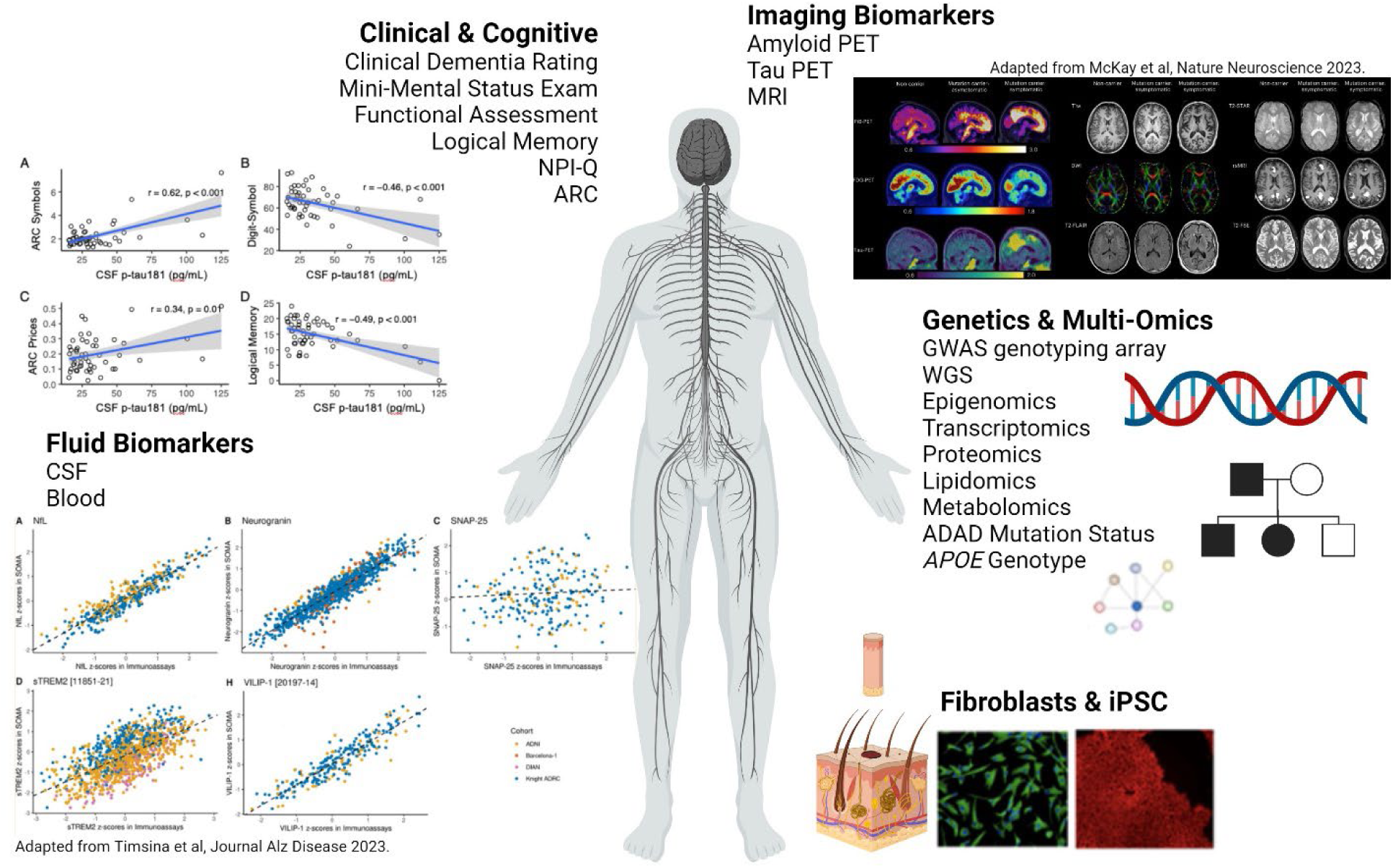
Overview of DIAN Obs Resource Requests

**Figure 5:**
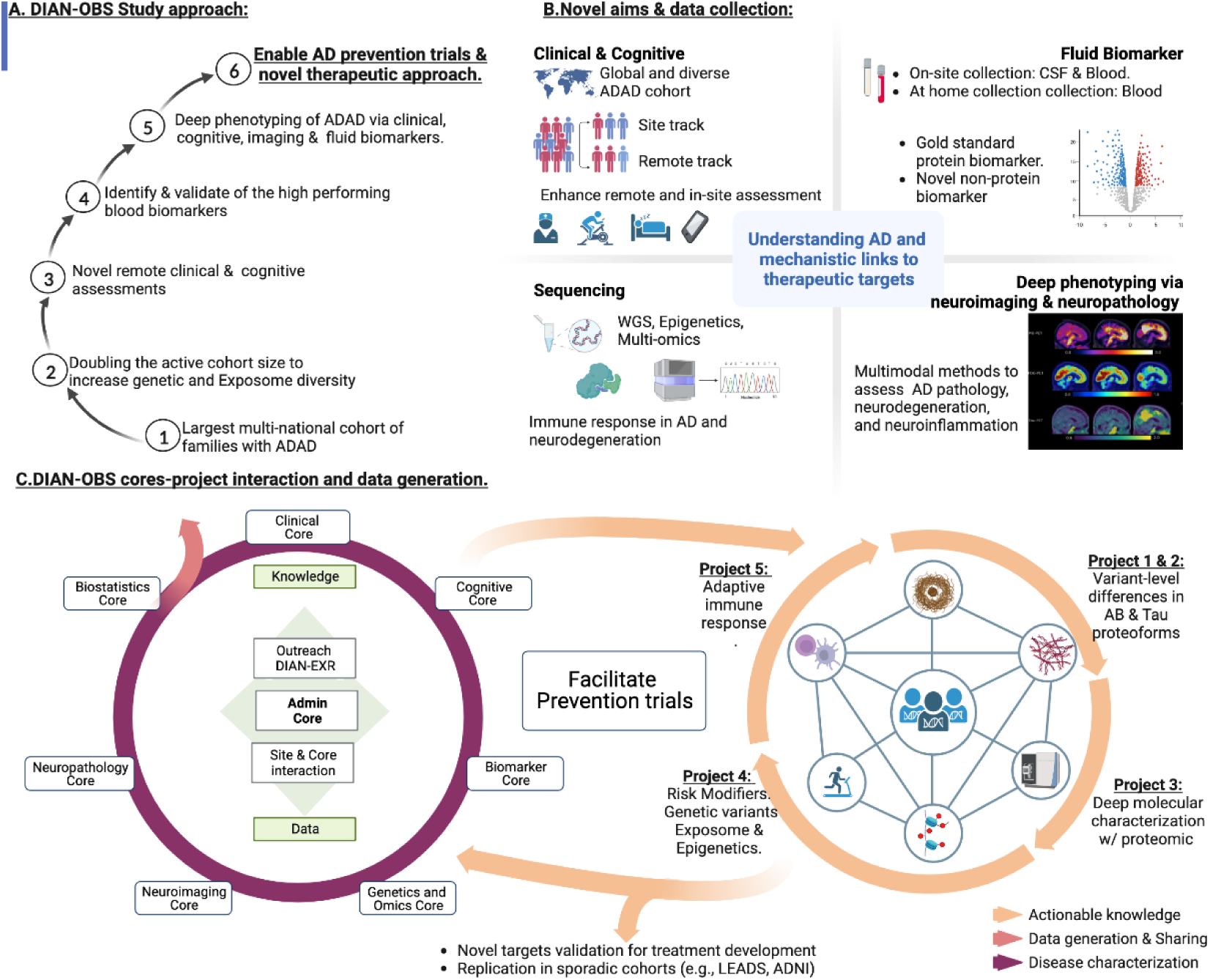
DIAN Obs Comprehensive Study Approach

### D. Clinical Core

The Clinical Core oversees clinical protocol execution and associated activities, encompassing participant recruitment, retention, clinical evaluations, CSF and blood sample acquisition, safety protocols, and quality control initiatives. Enrollment eligibility criteria include being 18 years of age or older and having a known pathogenic ADAD mutation in the family with a risk of inheriting the mutation. All DIAN Obs participants have access to genetic counseling and testing.

Upon participant consent, a visit anniversary date is established, initiating the DIAN Obs visit schedule with a frequency of one to three years. In-person site visits commence with the initial assessment and subsequently alternate with remote assessments for asymptomatic participants. Symptomatic participants undergo annual in-person evaluations under the current protocol. Table 1 details clinical and cognitive data collection procedures and data availability. A comprehensive list of current and former DIAN Obs performance sites is included as a Supplement.

**Table 1:**
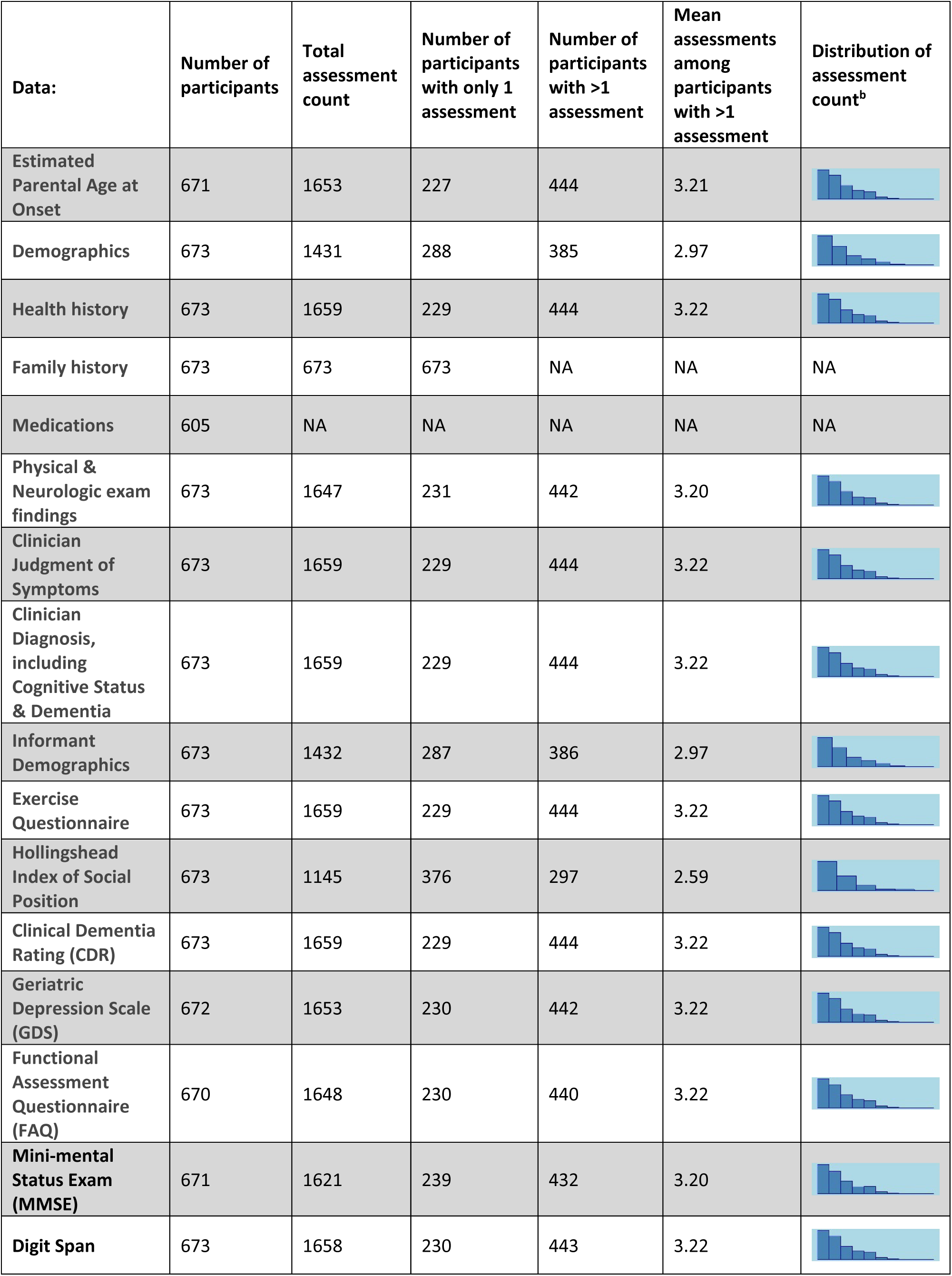
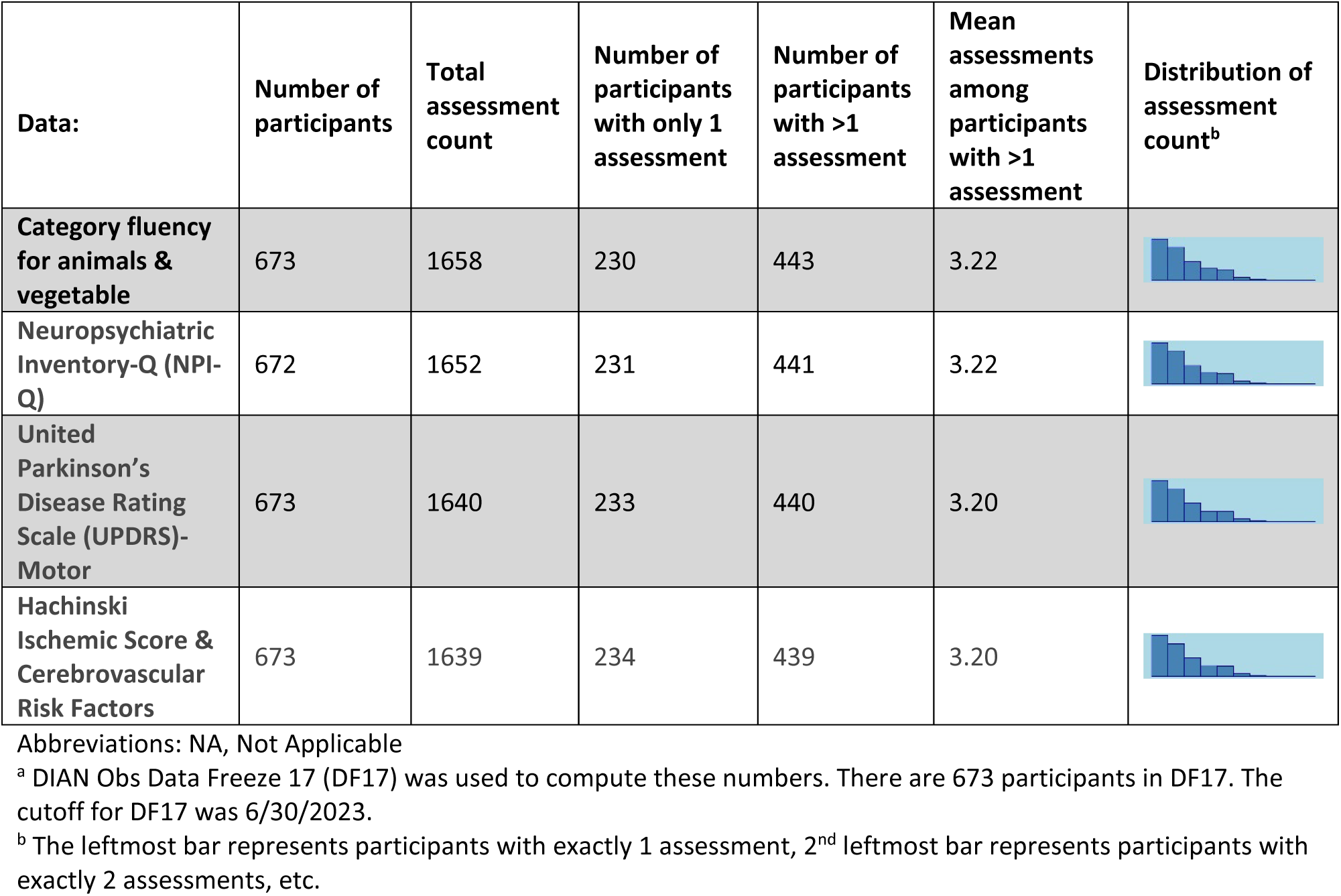
Clinical Core Available Resources & Data.

| **Data:** | **Number of participants** | **Total assessment count** | **Number of participants with only 1 assessment** | **Number of participants with >1 assessment** | **Mean assessments among participants with >1 assessment** | **Distribution of assessment count<sup>b</sup>** |
| --- | --- | --- | --- | --- | --- | --- |
| Estimated Parental Age at Onset | 671 | 1653 | 227 | 444 | 3.21 |  |
| Demographics | 673 | 1431 | 288 | 385 | 2.97 |  |
| Health history | 673 | 1659 | 229 | 444 | 3.22 |  |
| Family history | 673 | 673 | 673 | NA | NA | NA |
| Medications | 605 | NA | NA | NA | NA | NA |
| Physical & Neurologic exam findings | 673 | 1647 | 231 | 442 | 3.20 |  |
| Clinician Judgment of Symptoms | 673 | 1659 | 229 | 444 | 3.22 |  |
| Clinician Diagnosis, including Cognitive Status & Dementia | 673 | 1659 | 229 | 444 | 3.22 |  |
| Informant Demographics | 673 | 1432 | 287 | 386 | 2.97 |  |
| Exercise Questionnaire | 673 | 1659 | 229 | 444 | 3.22 |  |
| Hollingshead Index of Social Position | 673 | 1145 | 376 | 297 | 2.59 |  |
| Clinical Dementia Rating (CDR) | 673 | 1659 | 229 | 444 | 3.22 |  |
| Geriatric Depression Scale (GDS) | 672 | 1653 | 230 | 442 | 3.22 |  |
| Functional Assessment Questionnaire (FAQ) | 670 | 1648 | 230 | 440 | 3.22 |  |
| Mini-mental Status Exam (MMSE) | 671 | 1621 | 239 | 432 | 3.20 |  |
| Digit Span | 673 | 1658 | 230 | 443 | 3.22 |  |

| Data: | Number of participants | Total assessment count | Number of participants with only 1 assessment | Number of participants with >1 assessment | Mean assessments among participants with >1 assessment | Distribution of assessment count<sup>b</sup> |
| --- | --- | --- | --- | --- | --- | --- |
| Category fluency for animals & vegetable | 673 | 1658 | 230 | 443 | 3.22 |  |
| Neuropsychiatric Inventory-Q (NPI-Q) | 672 | 1652 | 231 | 441 | 3.22 |  |
| United Parkinson's Disease Rating Scale (UPDRS)-Motor | 673 | 1640 | 233 | 440 | 3.20 |  |
| Hachinski Ischemic Score & Cerebrovascular Risk Factors | 673 | 1639 | 234 | 439 | 3.20 |  |

Abbreviations: NA, Not Applicable
<sup>a</sup> DIAN Obs Data Freeze 17 (DF17) was used to compute these numbers. There are 673 participants in DF17. The cutoff for DF17 was 6/30/2023.
<sup>b</sup> The leftmost bar represents participants with exactly 1 assessment, 2<sup>nd</sup> leftmost bar represents participants with exactly 2 assessments, etc.

### E. Genetics Core

The goals of the Genetics Core are to obtain and bank tissue for genetic and multi-omic studies, as well as to generate, process, and/or harmonize genetic and multi-omic data. At each DIAN Obs in-person initial visit, whole blood is collected and used for ADAD mutation status, *APOE* genotype, genome-wide association study (GWAS) genotyping array, and whole genome sequencing (WGS). At each DIAN Obs in-person initial and follow up visit, buffy coat blood and PAXgene blood tubes are collected for longitudinal assessment of DNA methylomics and RNA transcriptomics, respectively. Additionally, the Core obtains and banks dermal fibroblasts then generates induced pluripotent stem cells (iPSCs) (Table 3). These resources support DIAN Obs Projects and are available to the research community with an approved data and/or tissue request.

The Genetics Core provides central determination and confirmation of gene sequence, whether normal or disease-causing mutation carrier, and *APOE* genotype on each of the 673 total study participants derived from 251 families. Table 2 presents the distribution of genetic variants and carrier status, categorized by gene. The Core, in coordination with the Clinical Core and DIAN EXR, maintains and curates a list of pathogenic mutations as well as confirms that new DIAN families carry an ADAD mutation.

**Table 2:** Genetics Core Available Resources & Data.

| **Data:** |
| --- |
| ADAD Mutation Status |
| APOE Genotype |
| Genome-wide association Study (GWAS) genotyping array |
| Whole Genome Sequencing (WGS) |
| Polygenic Risk Scores for: |
| ○ AD (risk, onset, progression), |
| ○ Parkinson's disease (risk) |
| ○ Frontotemporal dementia (risk) |
| Multi-omics: |
| ○ DNA methylomics: brain & blood |
| ○ RNA transcriptomics: brain & blood |
| ○ Proteomics: brain, CSF, & plasma |
| ○ Metabolomics & lipidomics: brain, CSF, & plasma |
| **Tissue & Cell Lines:** |
| DNA/Cell lines |
| Dermal Fibroblasts |
| Induced Pluripotent Stem Cells (iPSCs) |

**Table 3:** DIAN Obs Mutation Status Distribution.

| **PSEN1 (N = 436, 89 unique variants)** |  |
| --- | --- |
| mutation | 272 |
| no mutation | 164 |
| **PSEN2 (N = 50, 8 unique variants)** |  |
| mutation | 28 |
| no mutation | 22 |
| **APP (N = 120, 14 unique variants)** |  |
| mutation | 68 |
| no mutation | 52 |

The Genetics Core generates GWAS genotyping array, WGS, and *APOE* genotype data for all individuals. This data has been leveraged to generate polygenic risk scores (PRSs) for LOAD risk, onset, and progression, as well as Parkinson’s disease risk. Recent studies indicate that in DIAN Obs ADAD, LOAD risk PRS is not significantly associated with mutations status, but is associated with levels of CSF Aβ, total-tau, and p-tau, suggesting that known AD risk variants may modify age at onset (AAO) in the ADAD population <u>Polygenic risk score of sporadic late-onset Alzheimer’s disease reveals a shared architecture</u> <u>with the familial and early-onset forms - PubMed (nih.gov).</u>

Finally, the Genetics Core serves as a central repository for increasingly rich multi-omic data.^9,10^ Currently, this catalog includes: DNA methylomics (Illumina MethylationEPIC 850k array) for brain (44 participants) and buffy coat blood (790 longitudinal samples from 266 participants); RNA-seq for brain (44 participants) and blood (575 longitudinal samples from 319 participants); and proteomics (SomaLogic 7k) and metabolomics/lipidomics (Metabolon HD4) for brain (44 participants), CSF (495 participants), and plasma (495 participants). These data have been used to identify circular RNAs in brain associated with AD and AD pathology^41, 86^, as well as to identify proteins associated with carrying an ADAD mutation and change between 20 to −30 years before the onset, some of them even before some of the validated biomarkers.

### F. Cognition Core

A primary goal of the Core is to maintain the cognitive assessment battery to align with scientific aims and to incorporate novel measures and novel assessment methodologies that are more sensitive to early cognitive changes in ADAD. The Cognition Core serves the overall grant by overseeing rater training and maintaining rigorous quality control (QC) and documentation standards that ensure the fidelity of longitudinal cognitive assessments. In addition, the Cognition Core plays a pivotal role in maintaining the consistency of cognitive assessments across various languages, ensuring culturally relevant translations and adaptations across different sites and countries. These methodologies will improve reliability in measurement of the key features of ADAD. The assessment of cognition is central for achieving the scientific aims of all DIAN Obs Projects and Cores. The Cognition Core works with the Project and Core leaders to ensure that fully validated cognitive data is available for DIAN Obs data freezes and provide guidance on appropriate cognitive measures and data analyses to support Project and Core aims. Refer to Table 4 for Core data availability. Novel methods implemented in the Cognition Core include the use of remote cognitive testing via Ambulatory Research in Cognition (ARC), and the development of novel remote cognitive tasks including tests of long-term forgetting and statistical learning paradigms.

**Table 4:**
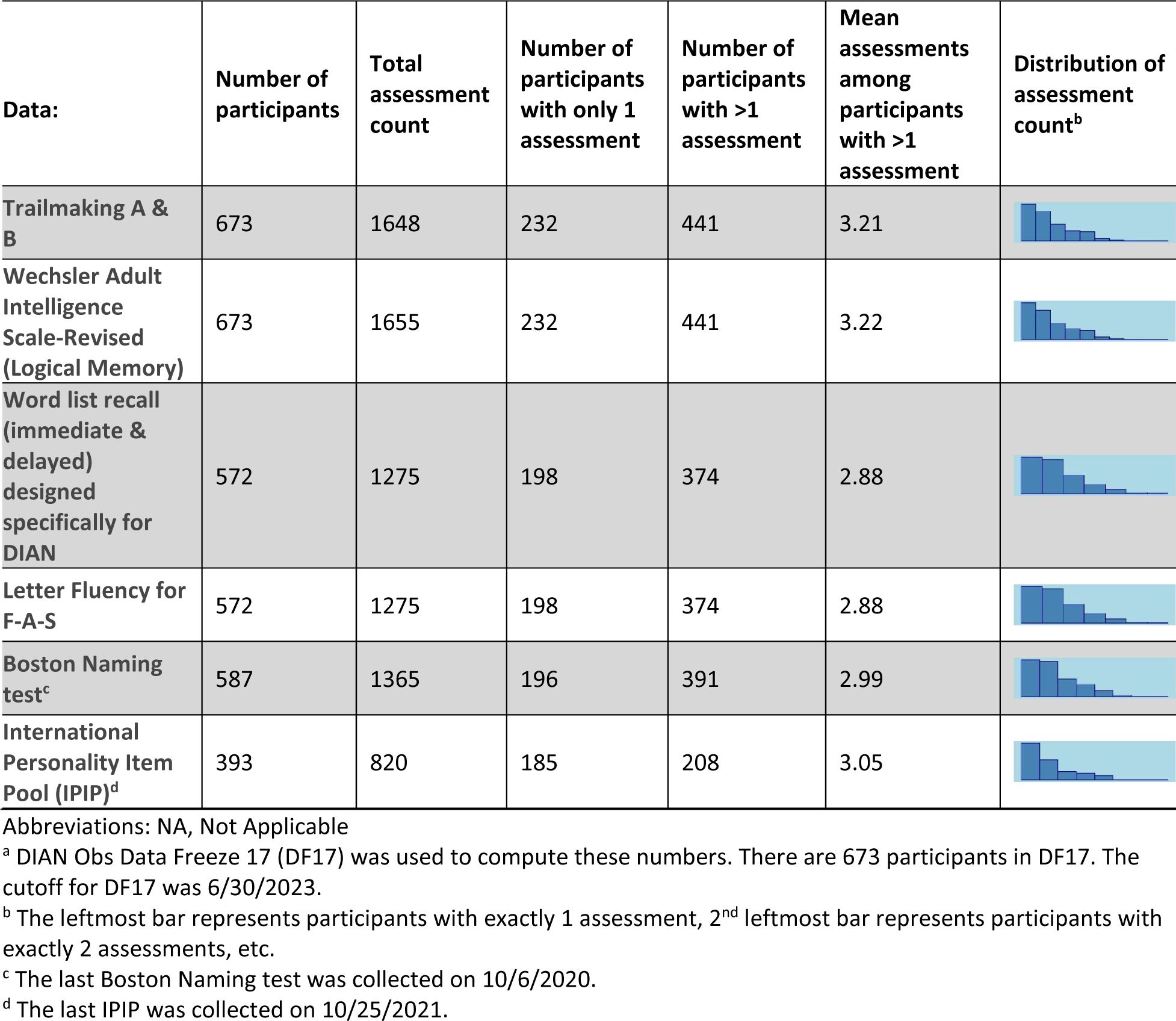
Cognition Core Available Resources & Data.

| **Data:** | **Number of participants** | **Total assessment count** | **Number of participants with only 1 assessment** | **Number of participants with >1 assessment** | **Mean assessments among participants with >1 assessment** | **Distribution of assessment count<sup>b</sup>** |
| --- | --- | --- | --- | --- | --- | --- |
| **Trailmaking A & B** | 673 | 1648 | 232 | 441 | 3.21 |  |
| **Wechsler Adult Intelligence Scale-Revised (Logical Memory)** | 673 | 1655 | 232 | 441 | 3.22 |  |
| **Word list recall (immediate & delayed) designed specifically for DIAN** | 572 | 1275 | 198 | 374 | 2.88 |  |
| **Letter Fluency for F-A-S** | 572 | 1275 | 198 | 374 | 2.88 |  |
| **Boston Naming test<sup>c</sup>** | 587 | 1365 | 196 | 391 | 2.99 |  |
| **International Personality Item Pool (IPIP)<sup>d</sup>** | 393 | 820 | 185 | 208 | 3.05 |  |

Abbreviations: NA, Not Applicable
<sup>a</sup> DIAN Obs Data Freeze 17 (DF17) was used to compute these numbers. There are 673 participants in DF17. The cutoff for DF17 was 6/30/2023.
<sup>b</sup> The leftmost bar represents participants with exactly 1 assessment, 2<sup>nd</sup> leftmost bar represents participants with exactly 2 assessments, etc.
<sup>c</sup> The last Boston Naming test was collected on 10/6/2020.
<sup>d</sup> The last IPIP was collected on 10/25/2021.

### G. Imaging Core

The Imaging Core is responsible for the acquisition, QC, processing, and analysis of the MRI and PET neuroimaging data for DIAN Obs. The imaging data set collected in DIAN Obs participants to date represents a highly valuable resource for AD research. It has supported cross sectional analysis of PET and MRI data to develop a timeline for imaging biomarkers in ADAD. Carriers of AD-causing mutations and their non-carrier (NC) siblings are enrolled and followed in the Clinical Core through the international DIAN Obs performance sites. Participants undergo structural and functional MRI, amyloid PET, tau PET, and metabolic PET imaging in conjunction with their clinical visits. The Core obtains and analyzes longitudinal imaging data that is fully integrated with clinical, psychometric, and CSF biomarkers, and allow for mutation-specific genotype-phenotype analysis. MRI data are processed using the Freesurfer Imaging suite to derive regions of interest. These regions are then used to process the PET data. Imaging Core data formats available are outlined in Table 5. A neuroimaging specific resource paper detailing in-depth imaging protocols has recently been published.^5,11^

**Table 5:**
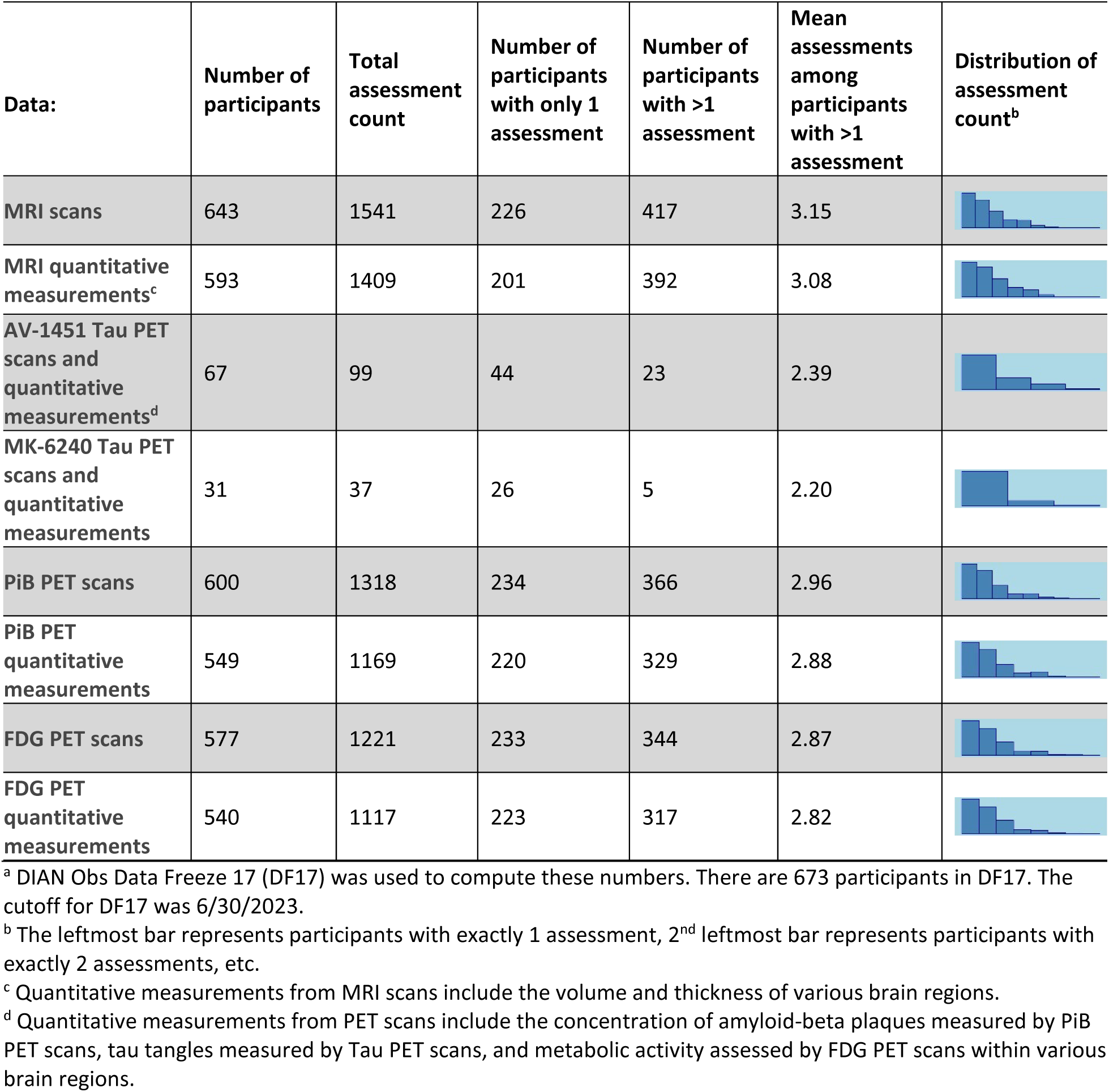
Imaging Core Available Resources & Data.

| **Data:** | **Number of participants** | **Total assessment count** | **Number of participants with only 1 assessment** | **Number of participants with >1 assessment** | **Mean assessments among participants with >1 assessment** | **Distribution of assessment count<sup>b</sup>** |
| --- | --- | --- | --- | --- | --- | --- |
| **MRI scans** | 643 | 1541 | 226 | 417 | 3.15 |  |
| **MRI quantitative measurements<sup>c</sup>** | 593 | 1409 | 201 | 392 | 3.08 |  |
| **AV-1451 Tau PET scans and quantitative measurements<sup>d</sup>** | 67 | 99 | 44 | 23 | 2.39 |  |
| **MK-6240 Tau PET scans and quantitative measurements** | 31 | 37 | 26 | 5 | 2.20 |  |
| **PiB PET scans** | 600 | 1318 | 234 | 366 | 2.96 |  |
| **PiB PET quantitative measurements** | 549 | 1169 | 220 | 329 | 2.88 |  |
| **FDG PET scans** | 577 | 1221 | 233 | 344 | 2.87 |  |
| **FDG PET quantitative measurements** | 540 | 1117 | 223 | 317 | 2.82 |  |

<sup>a</sup> DIAN Obs Data Freeze 17 (DF17) was used to compute these numbers. There are 673 participants in DF17. The cutoff for DF17 was 6/30/2023.
<sup>b</sup> The leftmost bar represents participants with exactly 1 assessment, 2<sup>nd</sup> leftmost bar represents participants with exactly 2 assessments, etc.
<sup>c</sup> Quantitative measurements from MRI scans include the volume and thickness of various brain regions.
<sup>d</sup> Quantitative measurements from PET scans include the concentration of amyloid-beta plaques measured by PiB PET scans, tau tangles measured by Tau PET scans, and metabolic activity assessed by FDG PET scans within various brain regions.

### H. Biomarker Core

The DIAN Obs Biomarker Core is a high-capacity biorepository enabling high-throughput processing while maintaining high-quality, gold-standard biomarker measurements of cerebrospinal fluid (CSF) and plasma samples available to investigators upon completion and approval through the DIAN tissue request process. The Biomarker Core obtains measures of the following biomarker analytes using the Lumipulse automated assay platform: CSF (Aβ1-40, Aβ1-42, total tau [t-tau], p-tau 181). Data and sample availability is outlined in Tables 6 and 7. Core samples may be leveraged in a longitudinal manner, in conjunction with extensive clinical and biological data, to study both traditional and exploratory biomarkers. With the main priority of the Biomarker Core to evaluate fluid biomarker profiles in ADAD participants comparing MCs to NC, DIAN Obs, along with others, have helped give insight to expected biomarker trajectories given the availability of expected AAO in MC individuals.^1,12^ The Biomarker Core has demonstrated that fluid biomarkers changes begin during the preclinical period (20-30 years before expected symptom onset). However, collection and analysis of additional longitudinal samples are required to define the patterns of change of known and novel fluid biomarkers that happen right when an individual progresses from asymptomatic (or preclinical) to symptomatic.

**Table 6:**
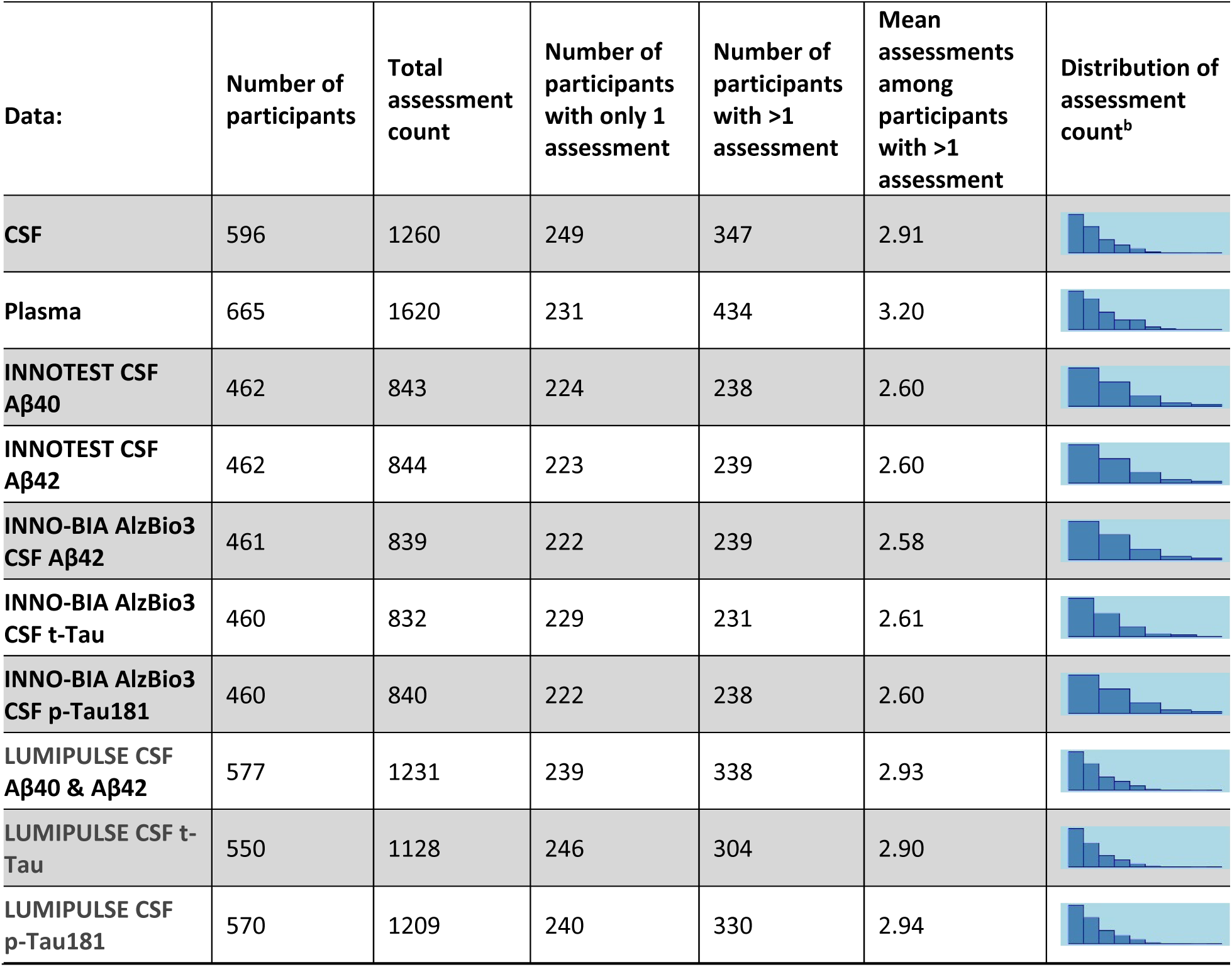
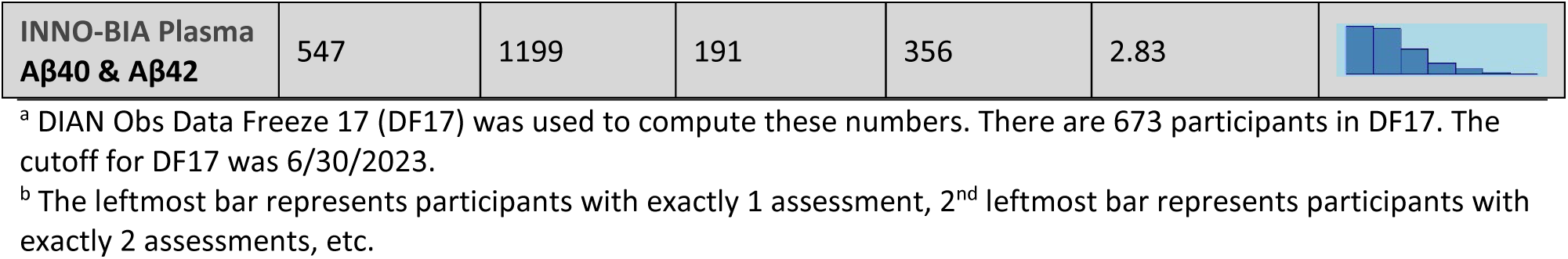
Biomarker Core Available Resources & Data.

| **Data:** | **Number of participants** | **Total assessment count** | **Number of participants with only 1 assessment** | **Number of participants with >1 assessment** | **Mean assessments among participants with >1 assessment** | **Distribution of assessment count<sup>b</sup>** |
| --- | --- | --- | --- | --- | --- | --- |
| **CSF** | 596 | 1260 | 249 | 347 | 2.91 |  |
| **Plasma** | 665 | 1620 | 231 | 434 | 3.20 |  |
| **INNOTEST CSF Aβ40** | 462 | 843 | 224 | 238 | 2.60 |  |
| **INNOTEST CSF Aβ42** | 462 | 844 | 223 | 239 | 2.60 |  |
| **INNO-BIA AlzBio3 CSF Aβ42** | 461 | 839 | 222 | 239 | 2.58 |  |
| **INNO-BIA AlzBio3 CSF t-Tau** | 460 | 832 | 229 | 231 | 2.61 |  |
| **INNO-BIA AlzBio3 CSF p-Tau181** | 460 | 840 | 222 | 238 | 2.60 |  |
| **LUMIPULSE CSF Aβ40 & Aβ42** | 577 | 1231 | 239 | 338 | 2.93 |  |
| **LUMIPULSE CSF t-Tau** | 550 | 1128 | 246 | 304 | 2.90 |  |
| **LUMIPULSE CSF p-Tau181** | 570 | 1209 | 240 | 330 | 2.94 |  |
| **INNO-BIA Plasma Aβ40 & Aβ42** | 547 | 1199 | 191 | 356 | 2.83 |  |

<sup>a</sup> DIAN Obs Data Freeze 17 (DF17) was used to compute these numbers. There are 673 participants in DF17. The cutoff for DF17 was 6/30/2023.
<sup>b</sup> The leftmost bar represents participants with exactly 1 assessment, 2<sup>nd</sup> leftmost bar represents participants with exactly 2 assessments, etc.

**Table 7:** Neuropathology Core Available Resources & Data.

| Data | Value | Frequency (Percentage) |
| --- | --- | --- |
| **A Score (Thal phase)** | 5 | 33 (100) |
| **B Score (Braak)** | 6 | 32 (97) |
| **B Score (Braak)** | 5 | 1 (3) |
| **C Score (CERAD)** | 3 | 33 (100) |
| **Sex** | Male | 17 (52) |
| **Race** | White | 28 (85) |
| **Race** | Other | 4 (12) |
| **Race** | Asian | 1 (3) |
| **Hispanic** | Yes | 4 (12) |

<sup>a</sup> DIAN Obs Data Freeze 17 (DF17) was used to compute these numbers. There are 673 participants and data from 33 brain autopsies in DF17. The cutoff for DF17 was 6/30/2023.

Given the minimally invasive nature of phlebotomy, the field is invested in the identification of plasma biomarkers. In response to the needs in the field, DIAN Obs’ scientific Projects will measure and analyze established markers of amyloid (Aβ and %p-tau217 ratio) and tau (MTBR243) deposition, inflammation (GFAP, sTREM2), and neurodegeneration (NfL) in CSF and plasma.

### I. Neuropathology Core

The Neuropathology Core houses the network’s post-mortem tissue. DIAN sites are supported in providing intact fixed hemi-brain specimens for uniform neuropathologic examination. Neuropathology Core efforts includes maintaining unfixed frozen and formalin-fixed tissue; a resource supporting DIAN’s Projects and available to the research community with an approved tissue request.

Fixed hemibrains are prepared in standard fashion (hemispheres coronally; cerebelli parasagittally; brainstems axially), digitally photographed, and sampled for histology, generating a set of 17 formalin-fixed, paraffin-embedded (FFPE) tissue blocks, representing the following areas: Middle frontal gyrus; anterior cingulate gyrus at the level of the genu of the corpus callosum; precentral gyrus; superior and middle temporal gyri; inferior parietal lobe (angular gyrus); occipital lobe (including the calcarine sulcus and peristriate cortex); posterior cingulate gyrus and precuneus at the level of the splenium; amygdala and entorhinal cortex; hippocampus and parahippocampal gyrus at the level of the lateral geniculate nucleus; striatum (caudate nucleus and putamen with nucleus accumbens) and olfactory cortex; lentiform nuclei (globus pallidus and putamen) at the level of the anterior commissure with the nucleus basalis of Meynert; thalamus with subthalamic nucleus; midbrain; pons; medulla oblongata; cerebellum with dentate nucleus; and cervical spinal cord. Remaining wet formalin-fixed tissue is kept in formalin in perpetuity as a research resource.

The Core prepares histology slides from a uniform set of seventeen FFPE blocks from each case. These are stained with hematoxylin and eosin for histomorphologic assessment, and with immunohistochemistry (IHC) for the more common neurodegenerative lesions, using antibodies for Aβ (10D5, Eli Lilly), phosphorylated tau (PHF1, Feinstein Institute for Medical Research, Manhasset, NY), α-synuclein (LB509,MilliporeSigma), and phosphorylated TAR DNA-binding protein of 43 kDa (pTDP-43, Cosmo Bio USA). This protocol enables the Neuropathology Core to identify and rigorously stage the pathological underpinnings of the major classes of neurodegenerative diseases. Histological slides are then reviewed and scored, using published semi-quantitative scoring criteria for histopathological lesions. These data inform formulation of diagnoses for each case (using consensus staging and neuropathological criteria for AD [Khachaturian, CERAD, NIA-Reagan Institute, and NIA-AA]^13–19^ and for non-AD disorders^20–28^). To date a total of 41 DIAN participant specimens have been secured. Core data and sample availability is outlined in Table 8.

### J. Biostatistics Core

The activities of the Biostatistics Core enhance the research objectives of DIAN by imparting a smooth transition from the database to statistical analyses, providing appropriate statistical analysis resources to all Cores and Projects, and developing longitudinal statistical models to test the preclinical hypotheses of DIAN on all major biomarkers of AD. The Core provides application of methodological significance as it is a necessity of state-of-the-art longitudinal statistical models to adequately estimate and compare the longitudinal rates of change on multi-modal biomarkers during the preclinical and symptomatic stages, and to assess their predictive power to cognitive decline.

The high dimensional data from Imaging Core from many modalities (MRI, PiB PET, Tau PET over a large number of brain regions) and the omics data from the Genetics and Multi-Omics Core present another unique analytic challenge to DIAN Obs. The Biostats Core seeks biologically meaningful dimension reduction, and conduct analyses to combine imaging markers and omics markers into composites for the test of critical hypotheses. Principal component analyses and partial least square analyses^29–30^ will be implemented, as well as methodologies developed by the Core^31–32^. We will also analyze longitudinal rates of change for these biomarkers jointly through general linear mixed models and correlate the rates of changes across modalities.

The Biostatistics Core have recently published multiple novel statistical methods driven by DIAN Obs database: analysis of biomarkers subject to detection limits^33^, correlations with family-clustered design^34^, diagnostic accuracy with ROC surface^35^, detection of unknown changepoints (in age) from multiple longitudinal biomarkers^36^, and a novel Bayesian ADAD progression model^37^. The Core continues to expand these models, and tackle other emerging analytic challenges from DIAN Obs: measurement errors in the EYO, small sample inferences on MCs who ‘escaped’ from their expected AAO, and high dimensional longitudinal data from imaging and omics. To control the false discovery rate (FDR), the Benjamini and Hochberg procedure is utilized^38^.

### K. Scientific Projects

In 2019, DIAN Obs added three scientific Projects to the study: Project 1: Amyloid-beta, Project 2: Tau, and Project 3: Novel Mechanisms. The goal of these scientific projects are to uniquely address central scientific questions which require significant DIAN Obs Core involvement.

Project 1 aimed to define the impact of ADAD mutations and amyloidosis on amyloid β proteoforms in CSF and plasma. This was accomplished using an IP-mass spectrometry approach to capture major Aβ proteoforms in plasma, CSF, and brain. Specifically we monitored Aβ37, 38, 39, 40, 42, and 43 in CSF, plasma, and brain homogenates and observed that Aβ isoform patterns of change differ. Additionally, Project 1 aimed to describe the impact of ADAD mutations in human iPSC-derived neurons and the relationship of brain proteoforms with histologic amyloid structure.

The next phase of this project will combine cell-based characterizations of individual ADAD variants with mass-spectrometry, IHC, and ELISA based measures of Aβ burden in brain parenchyma, cerebrovasculature, CSF, and blood from ADAD pathogenic variant carriers participating in DIAN Obs and DIAN-TU. This will provide a path toward the understanding of molecular composition and variant-level diversity of deposited and soluble Aβ species and compare these to the biochemical properties of each variant. These studies will offer a unique bench-to-bedside investigation of which types of Aβ are likely to be pathogenic, which are likely to deposit in brain and vessel walls, and how anti-amyloid therapies alter the balance of soluble Aβ species in the CNS and peripheral circulation.

The goal of Project 2 is to quantify the amount and regional distribution of tau pathology utilizing PET to illustrate differences between mutation carriers and non-carriers, investigate connections between tau pathology and other biomarkers as well as cognitive decline. The Project also aimed to validate the specificity and sensitivity of tau PET tracers (MK6240, AV1451, & PI2620) in postmortem tissue. Project 2 works to measure using mass spectrometry tau proteoforms in CSF, brain tissue, and iPSC-derived neurons, relating them to mutation status, EYO, AD biomarkers, and cognitive measures.

Initial work done within the Project used samples collected through 2017. During the study’s current grant cycle, analyses have expanded to CSF on samples collect since 2017. This expanded analysis included 411 total samples which captured 67 individuals with longitudinal visits. From these samples, the Project derived measures of p-tau phosphorylated at different sites. Also these sample were used to generate MTBR-tau243 data.

Project 3 aims to map molecular interactions providing greater explanation on how ADAD mutations, inflammation, synaptic function, and associated therapeutic targets may influence one another. The molecular profiling is completed by transcriptomics and mass spectrometry-based proteomics. Project 3 also explores defining profiles of targeted and novel fluid markers of neuroinflammation and injury to evaluate biomarker levels pre-clinically to progression to predict cognitive decline. Targeted inflammation markers in CSF include YKL-40, sTREM2, and progranulin (via immunoassay), and neuronal injury markers include CSF VILIP-1, neurogranin, SNAP-25, and NfL and plasma NfL.

### L. Future Initiatives

DIAN Obs has led major scientific advances in the understanding of AD stages, CSF and plasma biomarkers, mechanistic links to therapeutic targets, and enabled ground-breaking prevention and interventional trials. DIAN Obs has helped define the sequence, timing, and magnitude of longitudinal AD biomarker changes decades before symptoms begin. This work directly led to the development and implementation of primary and secondary prevention trials for ADAD and the validation of the amyloid-tau-neurodegeneration (ATN) criteria. DIAN Obs intends to build on these advances to further understand major contributors to disease progression, resilience, and heterogeneity, and target validation for future therapeutics.

As DIAN prepares for its next phase, study hypotheses will expand and move to be supported by home based remote assessments, smartphone-based applications, and wearable technologies. Home health nurses visits are being incorporated into the DIAN protocol allowing for clinical and cognitive assessments and biospecimen samples to be collected at visits occurring in years between a participant’s in-person visit. Additionally, the Cognition Core will extend the use the ARC smartphone application to include novel measures of long-term forgetting rates, which has been shown to be highly sensitive in preclinical ADAD. Another novel task is a measure of statistical learning that assesses evaluates learning rates over several consecutive days. Pilot data show that this is extremely sensitive to AD biomarkers in a sporadic AD population and is well tolerated by participants. Finally, DIAN will increase the generation and leverage of omic technologies to not only answer the main questions of the study but also generate new hypotheses and discover new biomarkers.

DIAN will continue its outreach to new families and regions of the world. The success demonstrated with DIAN’s South American performance sites has laid the groundwork to explore additional sites in Chile and Puerto Rico. There is a need to expand diversity in populations minimally represented in the DIAN cohort, with discussions initiated with potential collaborators in South Africa, Morocco, and Nigeria. There is also interest in re-establishing performance sites in the Western United States serving families previously identified in the region by former DIAN sites.

DIAN will maintain its lead role in defining the profiles of targeted and novel fluid markers of neuroinflammation and neuronal and synaptic injury over the course of the disease and evaluate the ability of biomarker levels at baseline, and longitudinal change over time, to predict cognitive decline. The landscape of biomarkers is expected to change rapidly due to amyloid removal treatments approaches. During the next years, DIAN plans to explore different omic layers to better characterize ADAD, but also to identify novel biomarkers.

Two new scientific projects are being planned to examine modifiers of risk (Project 4) and immune contributors (Project 5). The aims of Project 4 include defining the clinical and molecular profile of ADAD family members at the extremes of risk and protection, and evaluating the impact of genetic, non-genetic, and epigenetic factors on AAO and progression in ADAD families. Project 5 will examine T cells in relation to increased areas of tauopathy and neurodegeneration as well as changes in T cell populations, including Aβ- and Tau-specific T populations detection in the peripheral blood.

### Technical Validation

The undertaking of the longitudinal follow-ups on DIAN Obs participants over a wide spectrum of biomarkers requires working with diverse research teams and at the same time attaining a common goal by following a unified protocol and methodology. This demands additional effort on the statistical data QC. The Biostatistics Core implements cross-Cores and Projects data QC before formal statistical analyses are conducted. These QC procedures are in addition to the standard Core- and Project-specific QC, and crucial for safeguarding the validity of statistical inferences.

The Clinical and Cognition Cores have transitioned to electronic Clinical Outcome Assessments (eCOA) using a single vendor to: minimize burden on sites, follow 21 C.F.R. § 11 (Part 11) compliance more easily, and better harmonize the collection, storage and QC assessments of DIAN Obs. Contracted Clinical Research Associates (Monitors) perform site monitoring for compliance of the DIAN Obs protocol and accurate entry of data into the electronic data capture (EDC) system. The monitors track query resolution, enrollment, and help the Clinical Core track completion rates.

The collection, processing and storage of repository samples under Good Clinical Practices (GCP) using Part 11 compliant systems ensures the high quality of samples from which biomarker measurements will be available to approved investigators, we are enabling the scientific community to nominate and validate biomarkers in a highly phenotyped and homogeneous cohort.

Sites upload data to the DIAN Central Archive (DCA) managed by Flywheel. Scans undergo an initial automated QC of the DICOM for protocol deviations and PHI, then quarantined until cleared by PET University of Michigan) and MRI (Mayo Clinic) subcontractors. The University of Michigan reviews each PET for adherence to the specified tracer acquisition protocol including acquisition parameters, reconstruction method and parameters, smoothing kernel, application of image corrections (normalization, attenuation, scatter, randoms), pixel size and slice thickness, and upper and lower energy thresholds. For MRI, Mayo specialists review each session for protocol compliance and for image quality including intensity inhomogeneity and non-linearity issues.

Genetics Core QC procedures begin with receipt of samples when sample data is entered into the DIAN Obs database. Extracted DNA tubes are barcoded, and all subsequent DNA sample handling is tracked using barcoded tubes that are connected to the sample ID number. To avoid screening DNA samples for the wrong mutation, each site writes the familial mutation on the sample tube and on the family history form, as well as provides the Genetics Core with documentation from the physician or clinical laboratory that originally identified the mutation. If this information is missing or inconsistent, Genetics Core personnel contact the site coordinator to verify mutation information for the family. In addition to barcoded scanning and tracking we collect multiple samples from the same individual. Four tubes of whole blood are drawn at the participant initial visit, with two sent to the Genetics Core and two sent to National Centralized Repository for Alzheimer’s Disease (NCRAD at Indiana University School of Medicine). Whole blood and buffy coat blood samples received at the Genetics Core are derived from independent blood draws. ADAD mutation and *APOE* genotype are verified by comparing results across whole blood WGS and buffy coat blood whole exome sequencing (WES). Sample identity is verified by PLINK or KING genetic relatedness fingerprinting analysis of GWAS genotyping array, WGS, and WES. If fingerprinting data from these samples are concordant no further analyses are performed. If they are discordant we will DNA fingerprint the sample from NCRAD. Since NCRAD whole blood samples are drawn at the same time as the Genetics Core whole blood samples, they should be concordant unless a swap/mislabeling has occurred after blood draw. All the results from these QC procedures are recorded in the DIAN Obs database.

## Conclusions

The DIAN has provided seminal discoveries in AD pathophysiology and helped define the current understanding of the sequence of events that begin two decades before the first symptom onset, and progress through a decade of dementia. These advances are made on one of the world’s largest deeply phenotyped cohorts of both normal brain aging and AD progression. The data and sample sets are available to address questions and hypotheses on human brain function, aging, and AD, and with further utilization, promises to have even larger impacts.

## Supporting information

Supplementary Note 1 - DIAN Consortium Author List

## Data Availability

All data produced in the present study are available upon reasonable request to the authors. Requests may be submitted at: https://dian.wustl.edu/our-research/for-investigators/dian-observational-study-investigator-resources/

## Acknowledgements

Data collection and sharing for this project was supported by The Dominantly Inherited Alzheimer Network (DIAN, U19AG032438) funded by the National Institute on Aging (NIA), the Alzheimer’s Association (SG-20-690363-DIAN), the German Center for Neurodegenerative Diseases (DZNE), Raul Carrea Institute for Neurological Research (FLENI), Partial support by the Research and Development Grants for Dementia from Japan Agency for Medical Research and Development (AMED) (JP23dk0207066), the Korea Health Technology R&D Project through the Korea Health Industry Development Institute (KHIDI), Korea Dementia Research Center (KDRC), funded by the Ministry of Health & Welfare and Ministry of Science and ICT, Republic of Korea (HI21C0066), and Spanish Institute of Health Carlos III (ISCIII). This manuscript has been reviewed by DIAN Study investigators for scientific content and consistency of data interpretation with previous DIAN Study publications. We acknowledge the altruism of the participants and their families and contributions of the DIAN research and support staff at each of the participating sites for their contributions to this study. A full list of DIAN members appears in the Author Contributions.

## Supplement

**Supplementary Figure 1:**
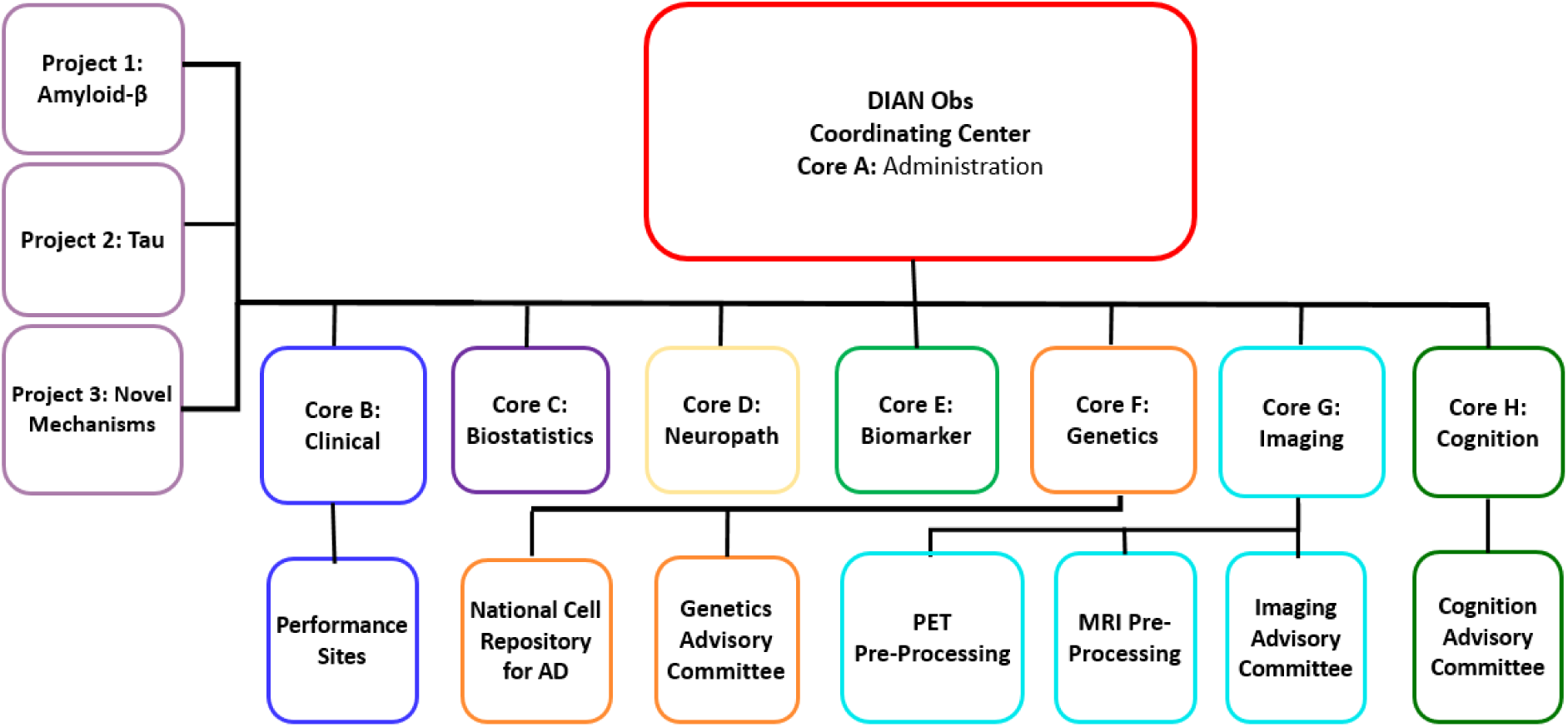
DIAN Obs Organizational Structure

**Supplementary Table 1.** DIAN Obs Clinical Sites (current & former)

| Site | Location | Site Leader | Activation Year | Status | # of Enrolled Participants |
| --- | --- | --- | --- | --- | --- |
| Washington University<sup>i</sup> | St. Louis, MO USA | Randall Bateman | 2008 | Current | 124 |
| University of California Los Angeles | Los Angeles, CA USA | John Ringman | 2008 | Former | 52 |
| University of Munich | Munich, Germany | Johannes Levin | 2013 | Current | 51 |
| University of Tübingen | Tübingen, Germany | Mathias Jucker | 2012 | Current | 45 |
| University College London | London, UK | Nicolas Fox | 2008 | Current | 41 |
| University of Pittsburgh | Pittsburgh, PA, USA | Sarah Berman | 2012 | Current | 40 |
| Indiana University | Indianapolis, IN USA | Martin Farlow | 2008 | Current | 39 |
| Edith Cowan University | Perth, Australia | Ralph Martins | 2008 | Current | 38 |
| Brigham & Women's Hospital | Boston, MA USA | Jasmeer Chhatwal | 2008 | Current | 35 |
| University of New South Wales | Sydney, Australia | Peter Schofield | 2008 | Current | 31 |
| Columbia University | New York, NY USA | James Noble | 2008 | Current | 30 |
| Butler Hospital | Providence, RI USA | John Huey | 2008 | Current | 28 |
| Grupo Neurociencias de Antioquia | Medellin, Colombia | Francisco Lopera | 2021 | Current | 27 |
| University of Melbourne | Melbourne, Australia | Colin Masters | 2008 | Former | 24 |
| FLENI<sup>ii</sup> | Buenos Aires, Argentina | Ricardo Allegri | 2015 | Current | 20 |
|  | Salta, Argentina | Patricio Chrem | 2021 | Current | 11 |
| Mayo Clinic | Jacksonville, FL USA | Gregory Day | 2013 | Current | 9 |
| Asan University<sup>iii</sup> | Seoul, South Korea | Jae-Hong Lee | 2015 | Current | 8 |
| University of Osaka | Osaka, Japan | Hiroshi Mori | 2017 | Former | 7 |
| Hirosaki City University | Hirosaki, Japan | Mikio Shoji | 2017 | Former | 5 |
| University of Tokyo | Tokyo, Japan | Yoshiki Niimi | 2017 | Current | 5 |
| University of Southern California | Los Angeles, CA USA | John Ringman | 2016 | Former | 4 |
| Niigata University | Niigata, Japan | Takeshi Ikeuchi | 2017 | Current | 4 |
| Hospital Clinic i Provincial | Barcelona, Spain | Raquel Sanchez-Valle | 2019 | Current | 2 |
| Instituto Nacional de Neuro | Mexico City, Mexico | Ana Luisa Sosa | 2023 | Current | 0 |
| University of Guadalajara | Guadalajara, Mexico | Victor Sanchez | 2023 | Pending | 0 |
| University de Sao Paulo | Sao Paulo, Brazil | Leonel Takada | 2023 | Pending | 0 |
| McGill University | Montreal, Canada | Pedro Neto-Rosa | 2023 | Current | 0 |
| **Total Enrolled:** |  |  |  |  | **673** |

i. Washington University in St. Louis is also the DIAN Coordinating Center.
ii. FLENI Salta satellite site
iii. Asan University reactivated in 2022.

**Supplementary Table 2.**
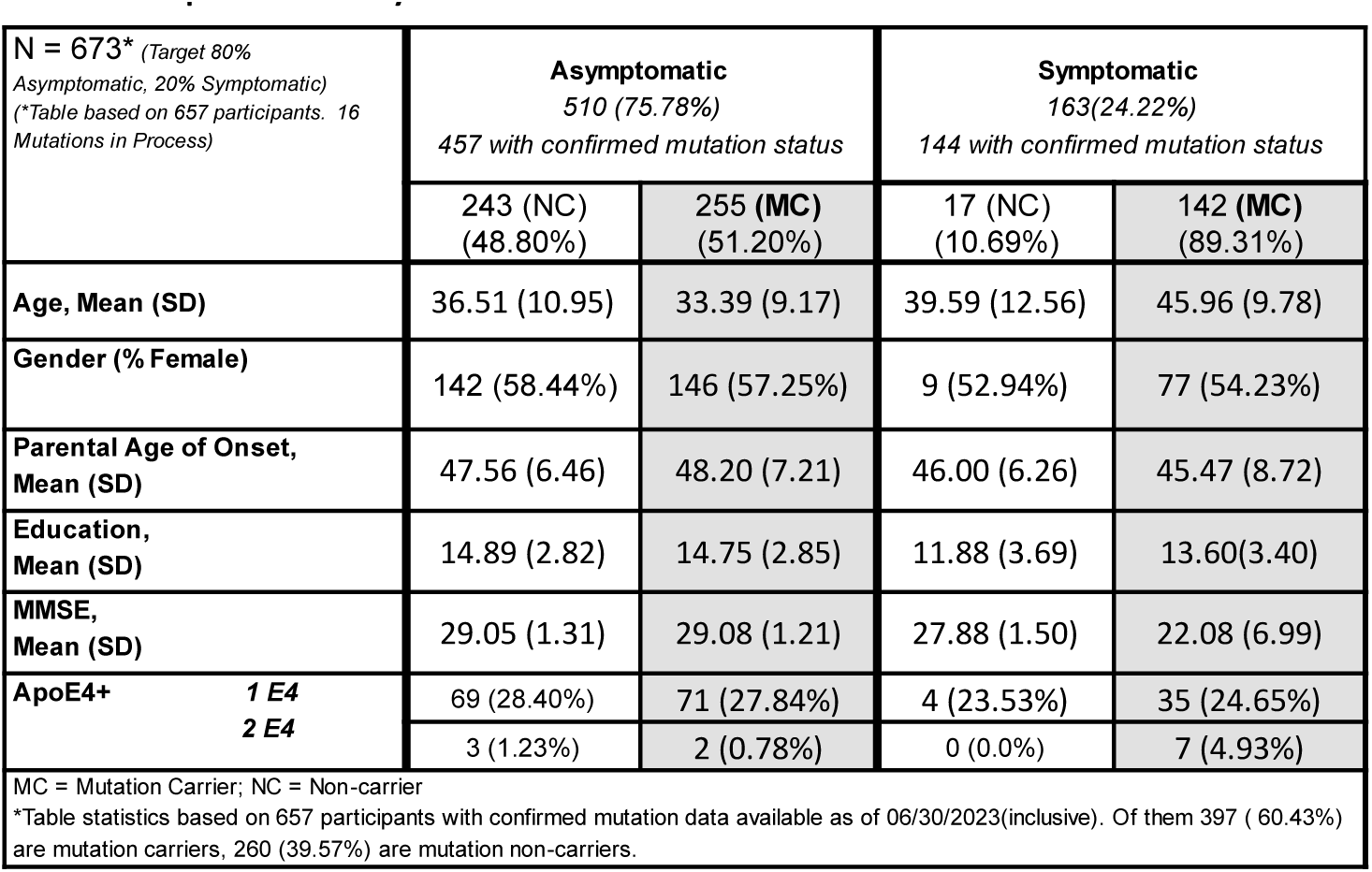
DIAN Obs Participant Entry Characteristics at Enrollment.

---

**Supplementary Note 1** – Author Contributions: DIAN Consortium List

Please see ‘DIAN Consortium Author List 2023.xlsx’ document.

